# Surveying Argentine healthcare providers’ knowledge, attitude, and practice about diagnosis and management of sports-related concussions

**DOI:** 10.1101/2023.06.20.23291643

**Authors:** María Julieta Russo, Fernando Salvat, Agostina Kañevsky, Marcelo Saco, Ignacio Alonso Hidalgo, Franco Della Vedova, Ricardo Francisco Allegri, Gustavo Sevlever

## Abstract

**Background:** A better understanding of the healthcare providers’ knowledge, attitude, and practice (KAP) level would help athletes, trainers, and athletic administrators, implement more effective concussion-management recommendations and guidelines. This survey aims to understand healthcare providers’ KAP dimensions regarding the diagnosis and management of sports-related concussions.

**Methods:** Cross-sectional study. An online-based KAP survey was carried out on a convenience sample of healthcare providers, coaches, parents, and athletes (n=626) in partnership with sports concussion clinics, rugby union, and rugby league in Argentina. The questionnaire incorporated 25 questions. Descriptive analysis was estimated as means, SD, and proportion. Chi-square tests, two-sample t-tests, and regression analysis were utilized for the response analysis.

**Results:** Seventy-four percent of the respondents reported having concussion training. Respondents correctly answered on average 6.23 ± 2.16 (out of 10) knowledge questions. The largest gaps were related to the clinical interpretation of symptom severity and neuroimaging. The smallest gaps were identified in young athletes’ management. There was a significant difference in mean survey respondents’ knowledge scores [F (1,622) =109.479, p<0.001] between those who had received formal concussion training and those who had not.

**Conclusions:** This study reveals that healthcare providers have appropriate knowledge and attitude regarding sport-related concussions but there are important knowledge gaps and practices that are often wanting. Our findings confirm the need for training and education on sport-related concussions. It would be advisable to implement educational campaigns specifically focused on the diagnosis and management of sports concussions, to raise awareness about this topic in schools, leagues, and sports associations.

## 1. INTRODUCTION

Concussions, a type of mild traumatic brain injury, are a frequent concern for those playing sports, especially contact sports. Athletes who have had a concussion have a greater chance of getting another concussion, and when a concussion is recurrent, athletes may be at risk of a wide range of short- or long-term complications. ^1, 2^ Early identification and proper management and treatment of a sport-related concussion is often a challenge and plays a crucial role in providing the best possible outcome.

Worldwide concussion legislation mandates that healthcare providers have experience not only in concussion diagnosis and management but also in the legal implications. For example, after the implementation of concussion legislation in all 50 United States Sports, the rates of treated concussions were higher than PR legislation trends.^3^ It is important to link knowledge transfer and action plans to achieve impact at individual, organizational, community, national, and international levels. Unfortunately, standards for permanent education on these issues are not yet standardized in all settings.

The care of athletes who have sustained a sports concussion is generally assigned to a team of sports medicine providers and healthcare professionals, which may include athletes, physicians, athletic trainers, sports neuropsychologists, physical therapists, managers, and/or referees. Regardless of discipline, the experience with and knowledge about sports concussions may be the most important factors for early recognition and effective concussion management, to reduce long-term sequelae.

A better understanding of the healthcare providers’ experience, beliefs, and knowledge level would help athletes, trainers, and athletic administrators, implement more effective concussion-management recommendations and guidelines. Therefore, the purpose of our study was to describe the current knowledge, attitude, and practice of Argentine healthcare providers in the diagnosis and management of sports-related concussions. In addition, we asked the respondents whether they had received formal concussion training through a formal class or course, and we analyzed the correlations between the formal training received and the variables of knowledge, attitude, and practice level. We hypothesized that the healthcare providers who had received formal training showed a better level of knowledge compared to those who had not.

## 2. METHODS

An online-based knowledge, attitudes, and practice survey was carried out on a convenience sample of healthcare providers, coaches, parents, and athletes in partnership with the Argentine rugby union and Sports Concussion Clinics in Argentina. The survey was conducted in July and August 2020 all over Buenos Aires, Argentina among healthcare professionals involved in the training and care of competitive or recreational athletes. Two physicians of the Sports Concussion Clinic (MJR and FS) announced and send out questionnaires by mailing lists. Participation was voluntary and anonymous. The survey sample consisted of people related to the health care of athletes at a recreational, amateur, elite, or professional level in Argentina (physicians, athletic trainers, sports neuropsychologists, physical therapists, managers, and referees). There were no incentives offered. Upon clicking on the submission button, it was checked whether the response was complete or not. The respondent was reminded and asked to answer in case any question was left unanswered. The whole questionnaire was a single page, the respondents replied by scrolling down to the next question. Each submitted response was checked for completeness. This functionality was available in the survey instruments by making all the questions mandatory. The respondents could review their answers before submission by scrolling up the page. The user could view the survey page only until s/he submitted the completed survey. The user was unable to view the survey again once it was completed and submitted.

### Survey development

We specifically designed for our study an online questionnaire with an accessible format to be completed by all participants. We conducted a pretesting survey of comprehension, acceptability, length and adherence, technical quality, and face and logical validity of the instrument by sending an e-mail invitation to selected physicians from our clinic, requesting each one to access the survey link, complete the questionnaire, and comment on these issues. The questionnaire was revised based on this first feedback response. We then discussed the revised questionnaire in three meetings.

The final version of the survey included a section on the socio-demographic characteristics of respondents; experience in sports; knowledge, attitude, and practice of Argentine healthcare providers about diagnosis and management of sports-related concussions.

Individual questions from previously published work addressing the level of knowledge, attitude, and practice on sports concussion ^4–9^ were modified for inclusion. Survey questions regarding attitudes and practice were measured on a 5-point Likert scale from strongly disagree to strongly agree. Sum scores were then calculated for all questions (max. 25 points). Knowledge questions were scored as correct and received one point or as incorrect and received zero points. Sum scores were then calculated for all questions (max. 10 points).

### Formal aspects

Approval from the ethics committee of the Ministry of Health of Buenos Aires, as well as the respective local ethic committee, has been obtained for this study. Participation in this survey was voluntary, and the confidentially of the respondents was preserved through all research stages and after. Survey respondents received information about the purpose of the research, the principal investigator, the length of time of the survey, and the confidentially in an information letter, which was sent together with the questionnaire. No personal information was collected or stored. All data were hosted in a project-specific site at the hospital server, with continuous Internet access. All data kept in the database were password protected. All data will be stored for at least five years after final publication.

Study design and reporting complied with the Checklist for Reporting Results of Internet E-Surveys (CHERRIES) ^10^.

### Statistical Analysis

The characteristics of survey respondents were analyzed with descriptive statistics (percentages and means ± standard deviations). Chi-square tests and a two-sample t-test were used to test the differences between those who had received formal concussion training and those who had not. To investigate whether concussion knowledge scores (dependent variable) varied by formal concussion training status (independent variable) after controlling for the effects of participant age and several years the participant reported having had experience in sport, an analysis of covariance (ANCOVA) was constructed. Graphical analysis of concussion knowledge scores indicated these data were normally distributed. The significance threshold for all analyses was set at p < 0.05, two-tailed. Statistical Package for Social Sciences (SPSS 21.0 for Windows, SPSS, Chicago, IL, USA) was used for all analyses.

## 3. RESULTS

A total of 626 participants completed and returned the questionnaire (response rate of 85%). After reviewing each questionnaire, we rejected 110 questionnaires because the answers were unclear, or the participant left unanswered items. The mean age of the participants was 42.49±12.49 years, and their average education was 15.93±3.43 years. Most participants were male (87.9%).

The sports experience of respondents is shown in Table 1. Their average experience in sports was 13±11.19 years. Most of the healthcare professionals were physicians (n=429; 68.5%). Nearly two-thirds (72%) of the respondents were related to rugby. Soccer was the second sport (6.2%). Seventy-four percent of the respondents (n=463) reported having formal concussion training (e.g., courses, talks, or specialization) in the management of sports-related concussions.

**Table 1.**
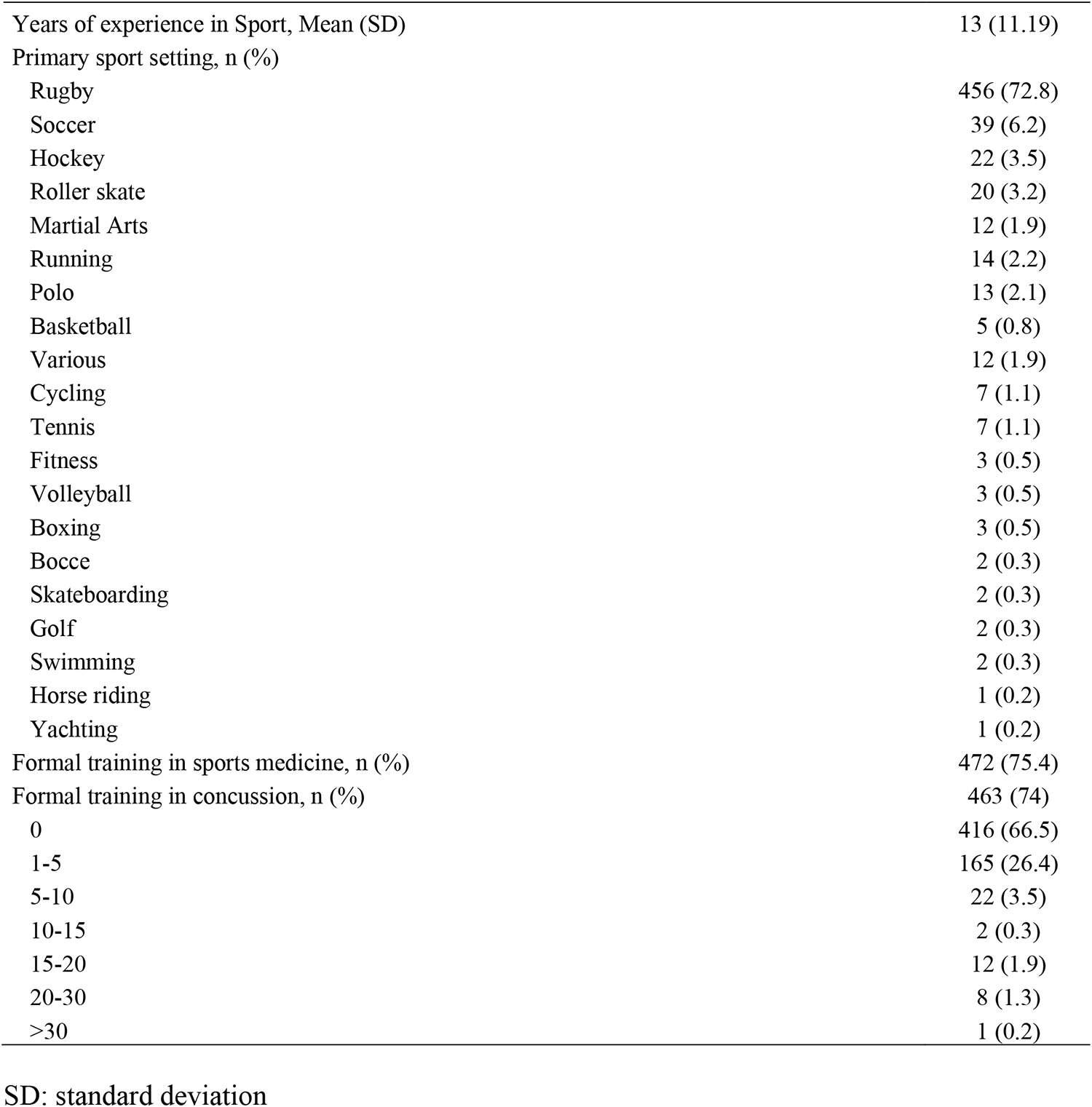
The sports experience of survey respondents.

The responses to the five questions regarding attitudes and practice about sports concussions are depicted in Fig. 1. Exactly, 63.6% of respondents agreed or strongly agreed that they are confident in their ability to identify a concussion, and almost 50% of respondents agreed or strongly agreed that they are confident in their ability to apply the decision-making process of returning an injured athlete to competition, and almost 85% of respondents agreed or strongly agreed that a multidisciplinary team with concussion experience should manage a person who has sustained a concussion and that concussions have a significant impact on the health of individuals and society. The distribution of responses to Q1-Q3 was similar across the five response categories. A higher rate of agreement was observed in Q4 and Q5 (Figure 1).

**Figure 1.**
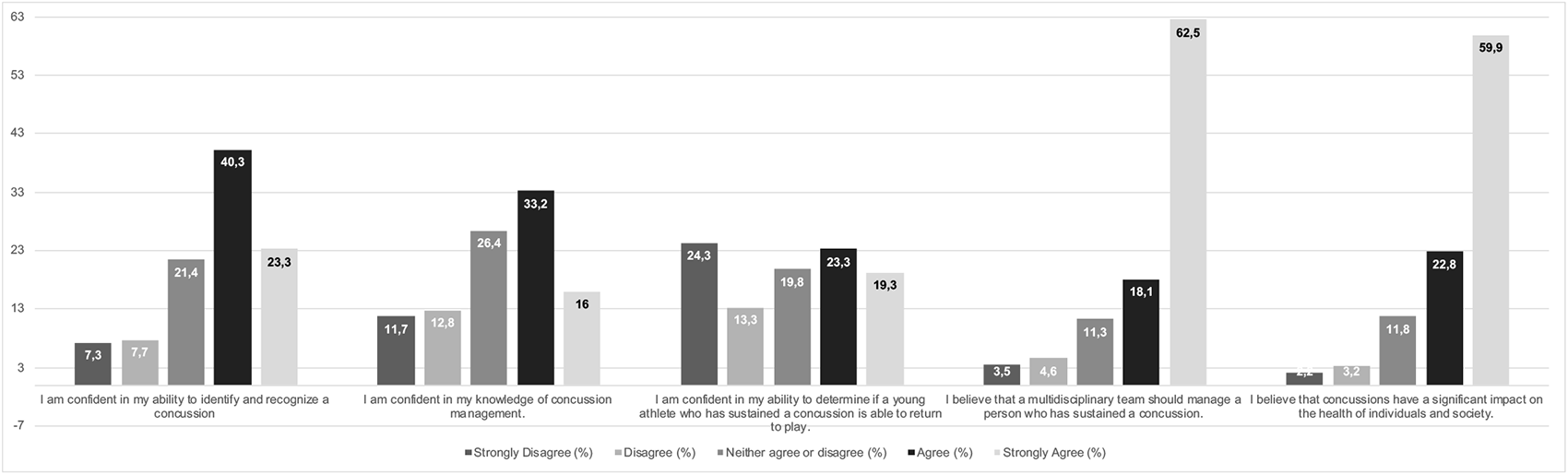
Attitudes and Beliefs of survey respondents.

Respondents correctly answered on average 6.23 ± 2.16 (out of 10) concussion knowledge questions. Seventy-four percent (n=463) of the respondents believed that neuroimaging (CT and/or MRI) is required to diagnose a concussion. Almost 60% answered false for the one true statement “a concussion is usually associated with a normal neuroimaging” and “a concussion sustained in youth should be managed more conservatively as a concussion sustained in adulthood”. Seventeen percent (n=105) answered true for the one false statement “Concussion management should be based on concussion severity grading scales”. The results of the 10 knowledge questions can be found in Table 2.

**Table 2.**
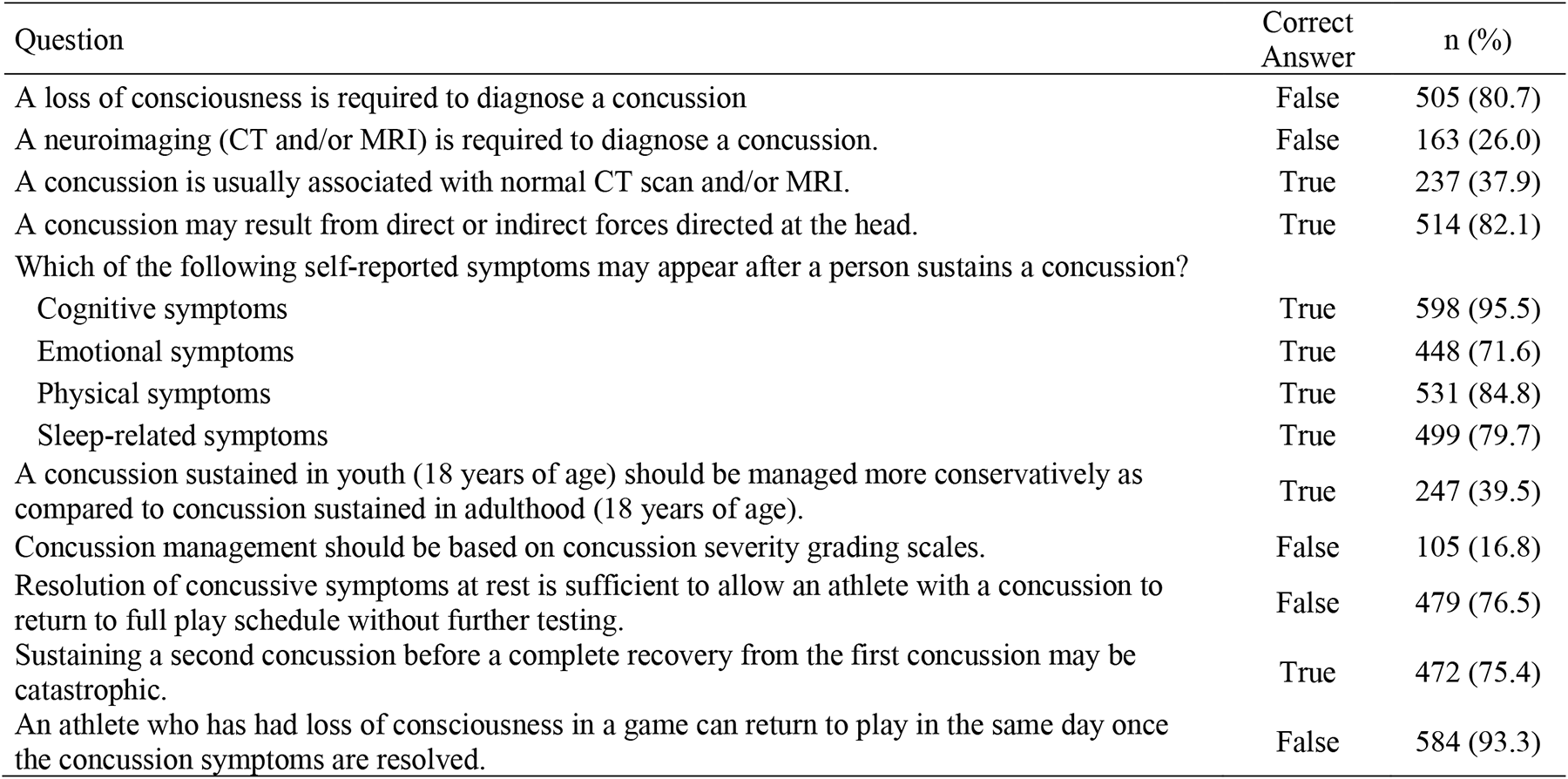
Percentages of correct answers for the knowledge questions.

| Question | Correct Answer | n (%) |
| --- | --- | --- |
| A loss of consciousness is required to diagnose a concussion | False | 505 (80.7) |
| A neuroimaging (CT and/or MRI) is required to diagnose a concussion. | False | 163 (26.0) |
| A concussion is usually associated with normal CT scan and/or MRI. | True | 237 (37.9) |
| A concussion may result from direct or indirect forces directed at the head. | True | 514 (82.1) |
| Which of the following self-reported symptoms may appear after a person sustains a concussion? |  |  |
| Cognitive symptoms | True | 598 (95.5) |
| Emotional symptoms | True | 448 (71.6) |
| Physical symptoms | True | 531 (84.8) |
| Sleep-related symptoms | True | 499 (79.7) |
| A concussion sustained in youth (18 years of age) should be managed more conservatively as compared to concussion sustained in adulthood (18 years of age). | True | 247 (39.5) |
| Concussion management should be based on concussion severity grading scales. | False | 105 (16.8) |
| Resolution of concussive symptoms at rest is sufficient to allow an athlete with a concussion to return to full play schedule without further testing. | False | 479 (76.5) |
| Sustaining a second concussion before a complete recovery from the first concussion may be catastrophic. | True | 472 (75.4) |
| An athlete who has had loss of consciousness in a game can return to play in the same day once the concussion symptoms are resolved. | False | 584 (93.3) |

A comparison of the survey results between those with and without formal training in concussion management is shown in Table 3. Both groups were closely matched in age, although respondents who had formal concussion training were more highly educated than respondents who had not trained. A greater proportion of respondents who had concussion training answered correctly both beliefs and knowledge questions.

**Table 3.** Comparison of the survey results between those with and without formal training in concussion management.

| Variables | Formal training in concussion management |  | t | p |
| --- | --- | --- | --- | --- |
|  | No | Yes |  |  |
| **Age (years)** | 41.26 (13.55) | 42.93 (12.08) | -1,468 | 0,143 |
| **Education (years)** | 15.34 (2.89) | 16.14 (3.58) | -2,593 | 0,01 |
| **Gender (male/female)** | 132/31 | 418/45 | 9,774 | 0,002 |
| **Years of experience in Sport** | 10.99 (11.92) | 13.71 (10.85) | -2,682 | 0,008 |
| **Total Beliefs score (out of 25)** | 16 (3.80) | 19.52 (3.36) | -10,452 | <0,001 |
| Q1: I am confident in my ability to identify and recognize a concussion | 2.72 (1.26) | 3.97 (0.88) | -13,721 | <0,001 |
| Q2: I am confident in my knowledge of concussion management. | 2.25 (1.16) | 3.66 (1.01) | -14,681 | <0,001 |
| Q3: I am confident in my ability to determine if a young athlete who has sustained a concussion is able to return to play | 2.24 (1.31) | 3.27 (1.41) | -8,185 | <0,001 |
| Q4: I believe that a multidisciplinary team should manage a person who has sustained a concussion | 4.5 (1.02) | 4.25 (1.08) | 2,564 | 0,011 |
| Q5: I believe that concussions have a significant impact on the health of individuals and society | 4.29 (1.06) | 4.37 (0.92) | -0,949 | 0,343 |
| **Total Knowledge score (out of 10)** | 4.82 (2.17) | 6.72 (1.93) | -10,463 | <0,001 |
| Q1: A loss of consciousness is required to diagnose a concussion. | 105 (20.80) | 400 (79.20) | 37,339 | <0,001 |
| Q2: A neuroimaging (CT and/or MRI) is required to diagnose a concussion. | 19 (11.70) | 144 (88.30) | 23,67 | <0,001 |
| Q3: A concussion is usually associated with normal CT scan and/or MRI. | 35 (14.80) | 202 (85.20) | 25,155 | <0,001 |
| Q4: A concussion may result from direct or indirect forces directed at the head. | 122 (23.70) | 392 (76.30) | 7,912 | 0,004 |
| Q5: Which of the following self-reported symptoms may appear after a person sustains a concussion? (Cognitive, emotional, physical, sleep-related symptoms) | 2.96 (1.05) | 3.44 (0.83) | -5,976 | <0,001 |
| Q6: A concussion sustained in youth (18 years of age) should be managed more conservatively as compared to concussion sustained in adulthood (18 years of age). | 39 (15.80) | 208 (84.20) | 22,252 | <0,001 |
| Q7: Concussion management should be based on concussion severity grading scales. | 79 (23.30) | 260 (76.70) | 2,871 | 0,055 |
| Q8: Resolution of concussive symptoms at rest is sufficient to allow an athlete with a concussion to return to full play schedule without further testing. | 86 (18.00) | 393 (82.00) | 69,224 | <0,001 |
| Q9: Sustaining a second concussion before a complete recovery from the first concussion may be catastrophic. | 97 (20.60) | 375 (79.40) | 30 | <0,001 |
| Q10: An athlete who has had loss of consciousness in a game can return to play in the same day once the concussion symptoms are resolved. | 136 (23.30) | 448 (76.70) | 34,197 | <0,001 |

There was a significant difference in mean survey respondents’ knowledge scores about concussion [F (1,622) =109.479, p<0.001] between those who had received the formal concussion training and those who had not, whilst adjusting for age and years the participant reported having had experience in the sport.

## 4. DISCUSSION

The identification of concussions remains an important task for healthcare providers. Although nearly all athletes make a full recovery after a sports concussion, if treated inappropriately, long-term complications can appear.^2, 11–13^ Therefore, it is imperative that all stakeholders comprehensively understand that to improve concussion care and athlete outcomes, standard recommendations must be disseminated and implemented at a national and local level and integrated into clinical practice. However, responsibilities and duties among professional groups and stakeholders vary across settings and individual levels of training and expertise. Adequate implementation of concussion care and management strategies should consider these issues.

This paper reports data from a cross-sectional survey on the current knowledge, attitude, and practice among healthcare providers in the diagnosis and management of sports-related concussion and their relationship to formal concussion training.

Most respondents agreed that a multidisciplinary team with experience should manage a person who has sustained a concussion and that concussions have a significant impact on the health of individuals and society. A similar pattern of responses was observed in physical therapists. ^8^ Healthcare providers must know that concussions should be properly diagnosed and managed to minimize potential negative effects. On the other hand, respondents demonstrated more confidence in recognizing a concussion (63.3%) than in determining a return to play (42.6%). Two previous studies reported similar findings.^8, 14^ Yorke et al. ^8^ mailed surveys designed to assess attitudes, beliefs, and knowledge of concussion to a sample of 55 physical therapists. While respondents correctly answered a large proportion of the concussion knowledge questions, they demonstrated less confidence to determine if an athlete who has sustained a concussion can return to play. Also, Carl et al. ^14^ found that pediatricians who reported managing at least one concussion per month were more comfortable with recognizing and managing concussions. On one hand, these reports seem to indicate that there is considerable knowledge about sports concussions, but on the other hand, there is an important need for more training on this topic.

In the present study, the findings indicated that people in our setting demonstrated good knowledge about sports concussions. However, misconceptions exist where 26% of the respondents believed that neuroimaging is required to diagnose a concussion or that a concussion is usually associated with abnormal neuroimaging (38%) and told that concussion management should be based on concussion severity grading scales (83%). In addition, the respondents showed limited knowledge of how sports concussions should be handled in young players whereas 60% said that a concussion sustained in youth should not be managed more conservatively than a concussion sustained in adulthood. Similar findings have been reported in previous studies. ^7, 15–18^

Although most respondents were physicians, they were less familiar with the more specific aspects of concussions, such as detection through computed tomography/magnetic resonance imaging or severity grading clinical scales. Similar results about whether a concussion could be identified with imaging were reported in other studies that surveyed hockey coaches ^7, 16^, soccer coaches ^15^, and physical therapists. Also, some authors have reported the misconception about the use of concussion severity scales to guide treatment.^8^ Based on current evidence, neuroimaging is not required to diagnose a concussion, and a concussion severity scale is not recommended to make return-to-play decisions ^19, 20^.

The current standard of treatment for sport-related concussion is a brief period of physical and cognitive rest (24–48 hours) until the athlete is asymptomatic, with a stepwise approach to rehabilitation before returning to sports participation. Of special concern is that the applicability of this protocol to children and adolescents is not clear. First, there are currently no suggested return-to-play protocol guidelines for youth athletes. ^19^ Second, youth athletes may report concussion symptoms differently from adults ^21^ and the clinical evaluation by the healthcare professional may need to include parent or teacher participation. Third, studies reveal a longer recovery period for youth athletes. ^22–24^ Lastly, youth athletes are one group with a higher risk of diffuse cerebral swelling because of a concussion ^25, 26^. Therefore, a more conservative approach to deciding when youth athletes can return to full sport is warranted. Our findings also revealed that many respondents did not know this issue related to younger athletes’ management.

The difference in answers between those who had received the formal concussion training and those who had not also was significant from a statistical point of view. Our results suggested that more trained healthcare providers rated their knowledge level as being significantly higher than those with less training, but significant findings did not differ after adjusting for age and years of experience in the sport.

In a systematic review analyzing the current evidence on sports concussion knowledge amongst sports coaches and match officials, ^27^ authors included ten studies assessing the level of prior education on sports concussion amongst coaches or officials. Studies suggest that the rate of formal concussion training is lower than what would be expected despite increasing public health concern.^4, 14, 28–35^. However, the influence of formal training on knowledge regarding concussions has been consistently shown by some authors. ^23, 29, 34, 36^

From a decision-making perspective, healthcare providers may benefit from specialized training focused on sports concussions. This suggests that formal concussion training will allow a greater knowledge and attitude transfer among people involved in contact sports, including players, parents, news media, coaches, referees, and healthcare providers.

As with other studies that employ survey research, this study was limited by several drawbacks. First, the sample size was also small and relied on convenience sampling in which specific rugby associations were asked to participate rather than random sampling. In some questions, there were too few subjects per category to provide meaningful results. Second, the lack of flexibility and potential depth may affect the validity of the results. Third, as with all studies relying on voluntary participation, results can be biased by a lack of respondents, if there are systematic differences between people who respond and people who do not. It is possible that participants who responded were more interested in concussions. Finally, a fixed-choice questionnaire might fail to explain the underlying reasons for the outcome.

## 5. PERSPECTIVE

Concussions in athletes are extremely common. The mismanagement of sports concussions can lead to long-term brain health consequences. This study revealed the knowledge, attitude, and practice of healthcare providers regarding the diagnosis and management of sports-related concussions, as such information was lacking in Argentina. Although healthcare providers have appropriate knowledge and attitude regarding sport-related concussions, there are important knowledge gaps and practices that are often wanted. Our findings confirm the need for training and education on sport-related concussions. It would be advisable to implement educational campaigns specifically focused on the diagnosis and management of sports concussions, to raise awareness about this topic in schools, leagues, and sports associations. Although it may be impossible to prevent all sports-related concussions, using survey-driven data as valuable evidence to influence the allocation of budgets, design of brain health education campaigns, and selection of content for training healthcare providers and athletes represents a desirable strategy to get more safe sports practices.

## Data Availability

All data produced in the present study are available upon reasonable request to the authors

## Acknowledgments

The authors are grateful to all survey respondents, athletes and families, rugby unions in Argentina, and Fleni Hospital.

## Ethical Statement

Ethical approval for this study was obtained from Fleni Institutional Review Board (APPROVAL NUMBER_14-18). Written informed consent was obtained from all subjects before the study.

